# Adolescent Weekend Catch-Up Sleep and Sleep Sufficiency: Protective Factors for Depression in Young Adulthood

**DOI:** 10.64898/2026.05.29.26354452

**Authors:** M. Pawley, S. Marwaha, B. I. Perry, I. Morales-Muñoz

## Abstract

**Background:** Sleep debt and irregular sleep patterns are highly prevalent amongst adolescents. However, whether the absence of these sleep behaviours protects against subsequent depression remains unclear. Here, we examined the association of sleep debt, weekend catch-up sleep (WCS), and social jetlag (SJL) in adolescence with depression in young adulthood and identified underlying biopsychosocial mechanisms.

**Methods:** Secondary data analyses were conducted using the Avon Longitudinal Study of Parents and Children. Bedtimes and wake-up times on school days and weekends (i.e., sleep duration) and sleep need were self-reported at 15 years. This was used to generate sleep debt (sleep need minus school day sleep duration), WCS (weekend sleep duration minus school day sleep duration), and SJL (absolute difference in the midpoint of sleep times between school days and weekends). Depression was assessed at 24 years with the Clinical Interview Schedule-Revised. Common mental health symptoms, biological, and school-related factors at 17 years were the mediators.

**Results:** Logistic regression analyses revealed that greater WCS (adjusted odds ratio [AOR]=0.90; 95% CI=0.84-0.97; *p*=0.004) and lower sleep debt (AOR=1.10; 95% confidence interval [CI]=1.03-1.18; *p*=0.005) at age 15 reduced the likelihood of depression at 24 years. Irritability at 17 years partially mediated the relationship between sleep debt and depression (bias-corrected estimate=0.003; 95% CI=0.002-0.004; *p*<0.001).

**Conclusions:** Adolescents who experience less sleep debt (i.e., less discrepancies between their actual sleep and their perceived sleep need) and those who extend their sleep duration on weekends are at reduced risk for depression in young adulthood. These findings underscore the need for more opportunities to increase adolescents’ hours of sleep to protect them against later poor mental health outcomes, such as depression.

**Key points and relevance:** *What’s known?:*

- Sleep debt and irregular sleep patterns are common in adolescence and have been linked with increased risk of depression; however, it remains unclear whether the absence of these sleep behaviours is protective and through which mechanisms this may occur.

*What’s new?:*

- Using a large, population-based cohort, we found greater weekend catch-up sleep and lower perceived sleep debt at 15 years to associate with reduced depression risk at 24 years, and irritability partially explained the latter relationship.

*What’s relevant?:*

- Regarding clinical practice, reducing adolescents’ perceived sleep debt and promoting longer weekend sleep duration may help lower subsequent depression risk. Additionally, delaying school start times may be an important education and public health policy target to support sufficient adolescent sleep.

## Introduction

Depression is a serious mental health problem in young people, with a rising global prevalence (Lu, Lin, & Su, 2024). Adolescence and early adulthood are periods of psychological vulnerability, with the peak onset for depressive disorders being 19.5 years (Solmi et al., 2022). Therefore, understanding the aetiology of depression in young people is crucial, and sleep has recently emerged as a relevant risk factor of interest (Scott, Kallestad, Vedaa, Sivertsen, & Etain, 2021). For instance, sufficient sleep quantity and quality is crucial during adolescence (Galván, 2020), and several studies have supported adolescents’ sleep problems as critical predicting factors for poor mental health (Scott et al., 2021). However, much of the research in young people has centred on specific and narrow aspects of sleep behaviour, particularly sleep duration (Becker, Sidol, Van Dyk, Epstein, & Beebe, 2017), without considering important intrinsic sleep characteristics.

Sleep consistency reflects the regularity of daily and/or weekly sleep-wake patterns and can be assessed using several dimensions of sleep (e.g., timings, duration, social jetlag) (Nicholson, Bohnert, & Crowley, 2023). Weekday-weekend sleep inconsistency (i.e., high variation between weekday and weekend sleep) increases until the end of adolescence (Nicholson et al., 2023). This may be attributed to adolescents’ circadian rhythm shift toward eveningness (Karan et al., 2021), which delays sleep-wake timings and creates a misalignment with early school start times (Crowley, Wolfson, Tarokh, & Carskadon, 2018). Furthermore, adolescents frequently report insufficient sleep duration on weekdays (Lemke et al., 2023), which may contribute to greater sleep debt by failing to meet their sleep need across the week (Guzzetti & Banks, 2023). To compensate for this, on the weekend adolescents often (1) extend their sleep duration (Nicholson et al., 2023); and (2) delay their bedtimes and wake-up times, creating a discrepancy between biological and social timing (Wittmann, Dinich, Merrow, & Roenneberg, 2006) (See Figure 1). This contributes to weekday-weekend sleep duration inconsistency (i.e., weekend catch-up sleep; WCS) and social jetlag (SJL), respectively, which along with sleep debt (Bakotic, Radosevic-Vidacek, & Koscec Bjelajac, 2017; Regestein et al., 2010) are associated with an increased risk of depression amongst adolescents (Lee, Kim, Jeon, Kim, & Park, 2022; Liu et al., 2020; Tamura & Okamura, 2024; Zhang et al., 2017). Nevertheless, whether the absence of these prevalent adolescent sleep behaviours reduces subsequent depression risk and which mechanisms are most relevant remains unclear. This may inform targeted intervention strategies to address rising global depression rates amongst adolescents (Lu et al., 2024).

**Figure 1.**
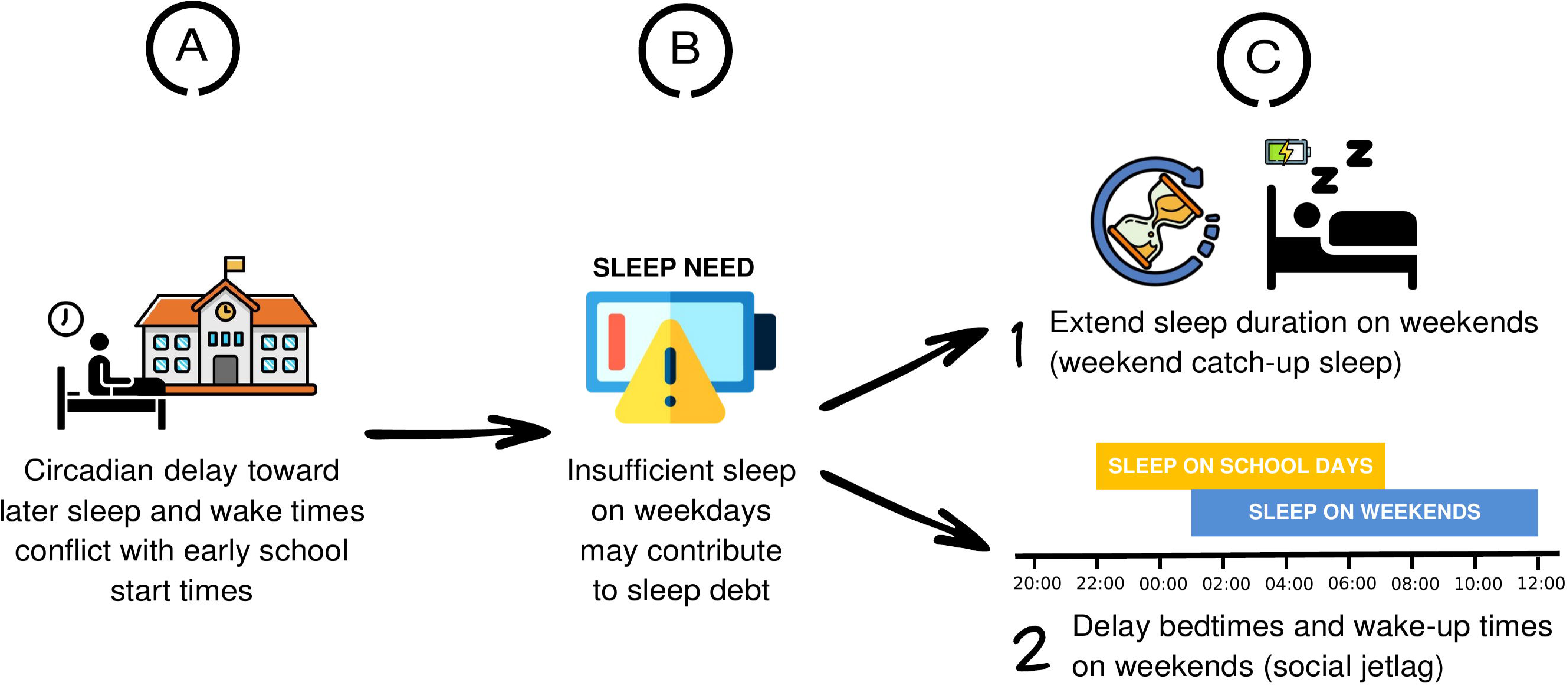

Several biopsychosocial factors have been proposed as potential underlying mechanisms between sleep and depression in young people; however, these have often been examined in isolation (Blake, Trinder, & Allen, 2018) which may overlook the complex multifaceted nature of depression. Examining factors in a holistic, integrated manner enables a deeper understanding of the aetiology of depression (Blake et al., 2018), which can inform effective preventative and treatment interventions that account for several interrelated and interacting risk factors (Marino et al., 2022).

The current study aimed to examine the prospective association of weekday-weekend sleep consistency, including WCS and SJL, and sleep debt in adolescence, with subsequent depression in early adulthood, and the potential mechanistic role of several biopsychosocial factors. Based on prior research, we hypothesised that non-normative adolescent sleep behaviours, specifically less WCS (Liu et al., 2020; Zhang et al., 2017), SJL (Tamura & Okamura, 2024), and sleep debt (Bakotic et al., 2017; Regestein et al., 2010), would decrease subsequent depression risk in young adulthood, and several biopsychosocial factors would mediate these associations.

## Methods

### Design and Setting

The Avon Longitudinal Study of Parents and Children (ALSPAC) is a longitudinal birth cohort study that recruited pregnant women based in Avon, the United Kingdom, with expected dates of delivery between 1^st^ of April 1991 and 31^st^ December 1992 (Boyd et al., 2013; Fraser et al., 2013). The initial number of pregnancies enrolled was 14,541, with a total of 14,676 foetuses. For this study, we used data from 3,039 participants comprising offspring who had attended the Teen Focus Clinic at 15 years, when sleep data was collected, and the Focus@24 years clinic (see Figure S1). Further information on the ALSPAC cohort is provided in Appendix S1. Details on all the data available can be found on the ALSPAC study website (http://www.bristol.ac.uk/alspac/researchers/our-data/).

Ethical approval for the study was obtained from the ALSPAC Ethics and Law Committee and local research ethics committees. Informed consent for the use of data collected via questionnaires and clinics was obtained from participants following the recommendations of the ALSPAC Ethics and Law Committee at the time.

### Measures

#### Sleep Behaviours in Adolescence

Adolescents completed a self-reported sleep questionnaire during the Teen Focus Clinic at 15 years. Questions that were relevant for this study assessed aspects of sleep separately on school days and weekends and were derived from a modified version of the School Sleep Habits Survey (Wolfson et al., 2003). From this, we generated three sleep predictor variables: weekend catch-up sleep (WCS), social jetlag (SJL), and sleep debt.

Sleep duration was assessed by calculating the difference between self-reported bedtime and wake-up time separately for school and weekend days. 19 adolescents with extreme sleep duration (<2 or >14 hours) were excluded from the analysis to control for potential inaccurate reporting. WCS was calculated as the difference between weekend and school sleep duration, mirroring other studies (Lin et al., 2018). Values that are zero reflect a consistent weekday-weekend sleep duration, positive values indicate longer weekend sleep duration compared to weekdays, and negative values reflect longer weekday sleep duration compared to weekend.

SJL represents a discrepancy between an individual’s social and biological clock due to misalignments in weekday-weekend sleep timings (Wittmann et al., 2006). This study utilised the sleep-corrected formula for social jetlag to assess the difference between the mid-point of sleep on weekdays and weekends whilst accounting for sleep debt (Jankowski, 2017). Higher scores indicate greater SJL (i.e., a larger mismatch in sleep/wake timings between weekdays and weekends).

Sleep debt was operationalised as the discrepancy between self-reported perceived sleep need and self-reported sleep duration on school days. Adolescents reported their perceived sleep need (i.e., the number of hours and minutes they felt they needed), which typically aligns with age-specific sleep duration recommendations (Godsell & White, 2019), supporting the use of this subjective measure as a reliable proxy for sleep need. Following prior research (Bakotic et al., 2017; Regestein et al., 2010), sleep debt was calculated by subtracting sleep duration on school days from sleep need, as adolescents typically accumulate sleep debt across the week and compensate by extending sleep duration on weekends (Nicholson et al., 2023). Higher scores indicate greater sleep debt.

#### Outcome

Depression diagnoses were assessed at age 24 using the Clinical Interview Schedule–Revised (CIS-R) (Lewis, Pelosi, Araya, & Dunn, 1992). The CIS-R assesses the severity, frequency, and persistence of various psychiatric symptoms to derive suggested diagnoses of common mental disorders in community samples, including depression. The CIS-R provides a diagnosis of depression according to the International Classification of Diseases– 10th Revision (ICD-10) criteria, including whether depression is mild, moderate, or severe. For this study, we utilised a binary variable to indicate the presence of depression through merging these three severity levels versus absence of depression (Perry et al., 2020).

#### Biopsychosocial Mediators

Mediators in this study reflect biopsychosocial characteristics assessed around 17 years of age that underlie the association between adolescent sleep and depression in young adulthood. Following methodological recommendations for longitudinal mediation (Li, Yoshida, Kaufman, & Mathur, 2023), we focused on factors measured at an intermediate time point (17 years) between the exposure (15 years) and outcome (24 years). We determined which factors to investigate based on a collaborative iterative approach using feedback from focus group discussions with the Youth Advisory Group (YAG) from the University of Birmingham’s Institute for Mental Health as well as existing scientific evidence. Appendix S2 and Tables S1 and S2 provide further information on this process and the supporting citations.

The final selected mediating factors included cardiometabolic function, school connectedness, perceived academic difficulties, fatigue, and irritability. Cardiometabolic function was proxied via measures of glucose-insulin homeostasis (the Homeostatic Model Assessment [HOMA_2_] for Insulin Resistance); and lipid homeostasis (total cholesterol [mmol/l]). School connectedness (Yes/No) was assessed with “How well do you feel that you fit in at your school or college?”. Perceived academic difficulties was determined by dichotomising responses (Yes/No) to “Do you have difficulty keeping up with your coursework or studies?”. Fatigue and irritability were separately derived from their corresponding section scores in the CIS-R (Lewis et al., 1992) which ranged in severity from zero (low) to four (high).

### Statistical analysis

A multi-staged analysis plan was developed. We first generated unadjusted and adjusted logistic regression models using SPSS (version 29) with all sleep variables at 15 years as predictors and depression at 24 years as the outcome. The adjusted (and primary) model controlled for confounders previously associated with depression: child’s gestational age (weeks) (Pyhälä et al., 2017), sex (male versus female) (Daly, 2022), ethnicity (white versus other ethnic minority) (Flores, Sharp, Carson, & Cook, 2023), prior depression (at 16 years; Short Mood and Feelings Questionnaire score) (Johnson, Dupuis, Piche, Clayborne, & Colman, 2018), early family adversity (Family Adversity Index score) (Wadman, Hiller, & St Clair, 2020), maternal age at birth (years) (Tearne et al., 2016), and concurrent sleep problems (yes versus no) (Chunnan, Shaomei, & Wannian, 2022). See Appendix S4 for further details. Odds ratios (ORs) and 95% confidence intervals (CIs) were generated. To handle missing data due to follow-up, we generated logistic regressions to identify factors associated with attrition (Table S3). Utilising these significant variables as predictors, a logistic regression model was conducted to generate weights for each individual using the inverse probability of response, following previous research (Hogan, Roy, & Korkontzelou, 2004; Kinner, Alati, Najman, & Williams, 2007). We used this weighting variable in the logistic regression analyses.

To examine the potential mediating role of the proposed biopsychosocial factors at age 17, we first tested whether these were associated with depression at 24 years and included all factors in the same model. An adjusted model was generated which incorporated the same covariates from the primary analysis. Factors that were significantly associated with depression were included as mediators in the path analysis between significant sleep predictors at 15 years from the primary model as exposures and depression at 24 years as the outcome using SPSS-AMOS (version 31). We conducted separate path analyses, one per sleep predictor. Given mediating factors could reflect either risk-enhancing or risk-reducing markers, we restricted the path models to sleep predictors and biopsychosocial mediators that associated with depression in the same direction (e.g., a risk-reducing sleep predictor was paired only with risk-reducing mediators, and vice versa). We controlled for the baseline covariates that were significantly associated with depression in the primary model and for the potential association between the biopsychosocial factors at 17 years. We used boot-strapped bias-corrected 95% CIs and *p* values to determine the significance of the standardised direct and indirect associations. Missing data were handled using the full information maximum likelihood method (Arbuckle, 1999).

## Results

The eligible sample for the primary analyses included 2,377 participants. The number of participants with missing data for each variable of interest is available in Table S4. Table 1 displays the frequency and descriptive values of all sleep, depression and covariate variables.

**Table 1.** Characteristics of the study population.

| Characteristic | N (%) | Range |
| --- | --- | --- |
| **Sleep predictor variables at 15 years** |  |  |
| Weekend catch-up sleep, mean (SD), hours (n=4734) | 1.53 (1.32) | -7.50 to 7.50<br>hours |
| Social jetlag, mean (SD), hours (n=4734) | 1.06 (0.80) | 0 to 7.00 hours |
| Sleep debt, mean (SD), hours (n=4651) | 0.38 (1.37) | -8.33 to 7.50<br>hours |
| **Depression outcome at 24 years** |  |  |
| Depression (n=3904) |  | NA |
| Yes | 422 (10.8) |  |
| No | 3482 (89.2) |  |
| **Covariates** |  |  |
| Child's gestational age, mean (SD), weeks (n=14363) | 38.41 (5.49) | 4 to 47 weeks |
| Maternal age when born, mean (SD), years (n=13835) | 27.97 (4.97) | 15 to 44 years |
| Early family adversity, mean (SD), FAI (n=11984) | 4.27 (4.44) | 0 to 32 |
| Depression at 16 years, mean (SD), SMFQ (n=4918) | 5.89 (5.62) | 0 to 26 |
| Ethnicity (n=11946) |  | NA |
| White | 11344 (95.0) |  |
| Non-white | 602 (5.0) |  |
| Sex (n=14773) |  | NA |
| Male | 7536 (51.0) |  |
| Female | 7237 (49.0) |  |
| Sleep problems at 24 years (n=3894) |  | NA |
| Present | 1450 (37.2) |  |
| No sleep problems | 2444 (62.8) |  |
| **Mediating factors at 17 years** |  |  |
| Glucose-insulin homeostasis, mean (SD), HOMA<sub>2</sub>, index (n=2923) | 0.97 (0.64) | 0.37 to 6.62 |
| Lipid homeostasis, mean (SD), Total cholesterol, mmol/L (n=3241) | 3.76 (0.68) | 1.44 to 7.33<br>mmol/l |
| School connectedness (n=3538) |  | NA |
| Yes | 3228 (91.2) |  |
| No | 310 (8.8) |  |
| Perceived academic difficulties (n=3562) |  | NA |
| Yes | 325 (9.1) |  |
| No | 3237 (90.9) |  |
| Fatigue, mean (SD), CIS-R subscale (n=4491) | 1.05 (1.43) | 0 to 4 |
| Irritability, mean (SD), CIS-R subscale (n=4491) | 0.89 (1.18) | 0 to 4 |

Note. Descriptive and frequency information for variables of interest among participants in the ALSPAC. N: Sample size; SD: Standard deviation; %: Proportion of the sample; NA: Not applicable; FAI: Family Adversity Index; SMFQ: Short Mood and Feelings Questionnaire; CIS-R: Clinical Interview Schedule – Revised.

Lower SJL (OR=1.212; 95% CI=1.107-1.327; *p*<0.001) and lower sleep debt (OR=1.116; 95% CI=1.054-1.183; *p*<0.001) at 15 years were associated with reduced risk for depression at age 24 in the unadjusted model (see Table 2 and Table S5). Furthermore, the association for sleep debt was consistent in the adjusted model (OR=1.104; 95% CI=1.030-1.184; *p*=0.005), but not for SJL (OR=0.933; 95% CI=0.827-1.053; *p*=0.263). WCS also emerged as significant (OR=0.899; 95% CI=0.835-0.967; *p*=0.004) in the adjusted model, whereby greater WCS was associated with lower odds of depression.

**Table 2.** Weighted unadjusted and adjusted associations between sleep variables at 15 and depression at 24.

|  | Unadjusted Model (N=2377) |  |  | Adjusted Model (N=1899) |  |  |
| --- | --- | --- | --- | --- | --- | --- |
|  | OR | 95% CI | *P* value | OR | 95% CI | *P* value |
| Weekend catch-up sleep | 0.968 | 0.909, 1.030 | .302 | 0.899 | 0.835, 0.967 | .004 |
| Social jetlag | 1.212 | 1.107, 1.327 | <.001 | 0.933 | 0.827, 1.053 | .263 |
| Sleep debt | 1.116 | 1.054, 1.183 | <.001 | 1.104 | 1.030, 1.184 | .005 |

Note. Logistic regression models included (unadjusted model) no potential confounders and (adjusted model) potential confounders including child's gestational age (weeks), sex (male/female), ethnicity (white/non-white), prior depression (at 16 years; Short Mood and Feelings Questionnaire score), early family adversity (Family Adversity Index score), maternal age at birth (years), and concurrent sleep problems (yes/no). Results for all confounders can be found in Supplemental Table 5. OR: Odds ratio; CI: Confidence interval.

To determine which mediators to include in the path analysis, adjusted regression analyses were generated to identify biopsychosocial factors significantly related to depression at 24 years (see Table S6). Irritability (OR=1.317; 95% CI=1.166-1.489; *p*<0.001), cholesterol (OR=1.512; 95% CI=1.231-1.857; *p*<0.001), and perceived academic difficulties (OR=1.742; 95% CI=1.181-2.568; *p<*0.001) were positively associated with depression (i.e., risk-enhancing factors), thus these were then selected as mediating factors for the path analyses. As sleep debt was similarly identified as a risk-enhancing marker for depression, a path analysis was run with this variable as a predictor. Furthermore, given early family adversity, maternal age at birth, and prior depression were significant baseline covariates in the primary model, these were controlled for in the path analyses.

In examining whether perceived academic difficulties, irritability, and cholesterol at 17 years partially mediated the relationship between sleep debt at 15 years and depression at 24 years, path analysis model fit indexes initially indicated poor model fit (χ^2^=959.970; *p*<0.001; root mean square error of approximation [RMSEA]=0.066; comparative fit index [CFI]=0.892). Inspection of the modification indices and standardised residual covariances highlighted previously unmodelled associations, therefore we sequentially included empirically supported direct paths (López-López et al., 2021; Moreno-Giménez et al., 2022; Parekh, Smeeth, Milner, & Thure, 2017; Riem & Karreman, 2019; Savage et al., 2015; Villadsen et al., 2023) until reaching good model fit (χ^2^=6.258; *p*=0.100; RMSEA=0.008; CFI=0.999). The final model included additional direct paths between depression at 16 years with all biopsychosocial mediators, early family adversity and perceived academic difficulties and cholesterol, as well as between maternal age at birth and irritability. Full details of the model re-specification process are provided in Appendix S5 and Table S7. Figure 2 presents the direct associations. Irritability mediated the relationship between sleep debt at 15 years and depression at 24 years (bias-corrected estimate=0.003; 95% CI=0.002-0.004; *p*<0.001), while perceived academic difficulties and cholesterol were not significant mediators in the exposure-outcome relationship.

**Figure 2.**
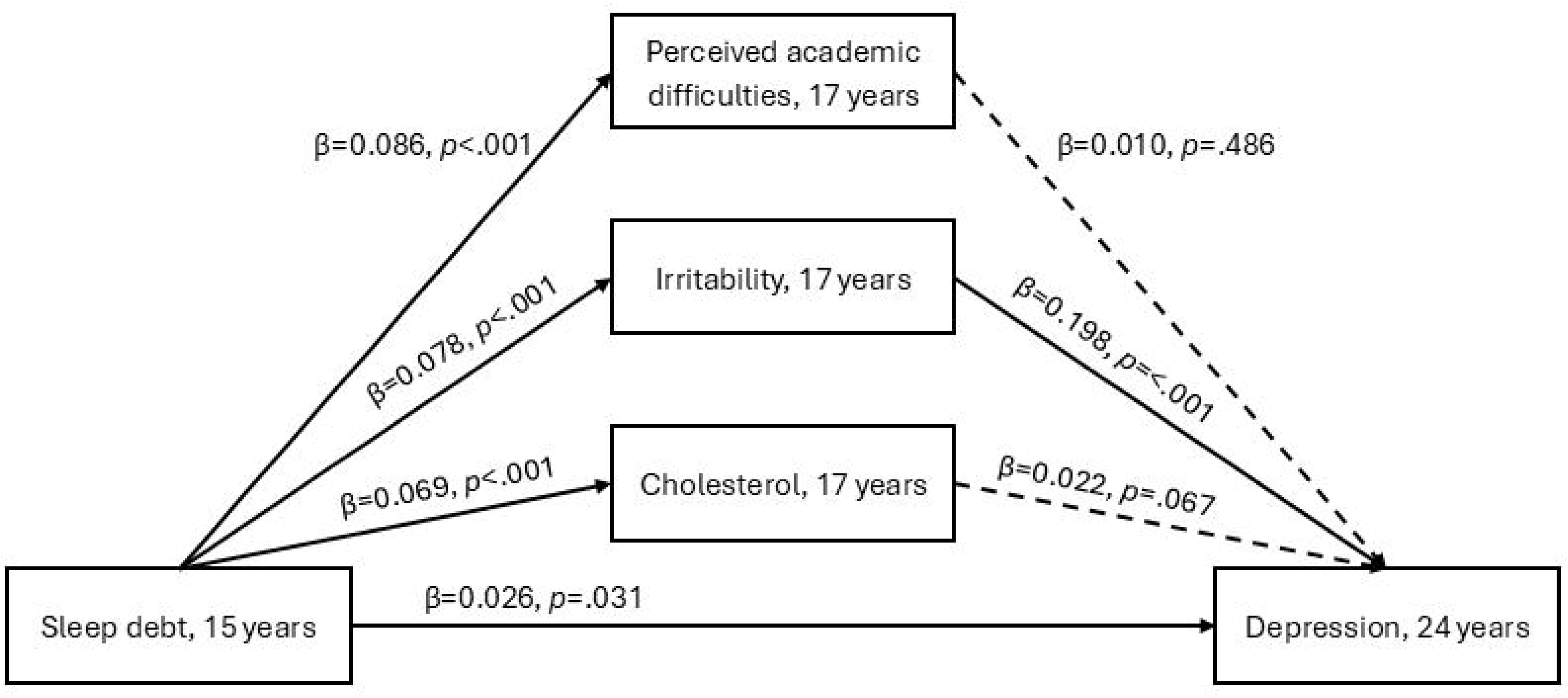

## Discussion

To our knowledge, this is the first study to report that greater WCS and lower perceived sleep debt at 15 years are associated with reduced depression risk in young adulthood, and that certain biopsychosocial mechanisms at 17 years underlie some of these relationships. Specifically, irritability partially mediated the association between sleep debt and depression. These results highlight the potential value of extending sleep duration on free days (i.e., weekends) as well as ensuring that the sleep needs are met during adolescence to help prevent future depression outcomes.

We first found greater WCS in adolescence to associate with lower depression risk in young adulthood. Adolescents often extend their sleep duration on weekends to recover from accumulated weekday sleep debt (Nicholson et al., 2023), which may have implications on subsequent mood and psychological functioning. Several cross-sectional studies have investigated the role of WCS on depression in adolescence, finding that greater WCS reduces (Koo et al., 2021) and increases (Lee et al., 2022; Zhang et al., 2017) risk, as well as no WCS (i.e., sleeping the same amount or more on weekdays than weekends) to positively associate with depression (Lee et al., 2022). This U-shape relationship has also been supported longitudinally whereby ≥3.5-hours of WCS and <0 hours (i.e., sleeping longer on weekdays than weekends) increased the risk of depressive symptoms one-to-two years later (Liu et al., 2020). These mixed findings cloud whether WCS confers risk or protection, or when such effects may manifest given the predominance of cross-sectional studies. Nevertheless, our results support that, over extended follow-up, greater WCS is associated with reduced depression risk. Further research is needed to determine the optimal range of WCS that minimises risk of subsequent depression.

Our findings demonstrated that lower perceived sleep debt at 15 years was associated with lower depression risk at 24 years. Prior cross-sectional (Bakotic et al., 2017; Regestein et al., 2010) and ecological momentary assessment (Shen, Wiley, & Bei, 2021) studies have largely focused on the finding that greater perceived sleep debt heightens depressive symptoms and next-day negative affect amongst youth. Experimental research further highlights partial sleep restriction over several nights to worsen mood and impair adolescents’ ability to regulate negative emotions (Baum et al., 2014). Our results likewise support greater sleep debt to increase depression risk; however, this can be interpreted from the complementary perspective whereby less sleep debt is linked to lower depression risk. Specifically, reducing the discrepancy between adolescent’s actual sleep and their sleep need may function as a protective factor for future depression, rather than solely conceptualising sleep debt as a vulnerability factor.

We additionally tested for biopsychosocial mediators in the prospective association between perceived sleep debt and depression, whereby irritability emerged as a significant mechanism. Adolescents with experimentally induced chronic partial sleep restriction report greater irritability (Baum et al., 2014) which is associated with depression in early adulthood (Hawes et al., 2020). Irritability is a transdiagnostic symptom representing emotion dysregulation across several emotional and behavioural disorders in childhood and adolescence (Chin, Robson, Woodbridge, & Hawes, 2025). Given both sleep (debt) and irritability are modifiable risk factors for depression (Mohamed, Croarkin, Jha, & Vande Voort, 2023; Short, Booth, Omar, Ostlundh, & Arora, 2020), ensuring adolescents meet their sleep need and can effectively manage their irritability through emotion regulation or stress management strategies may provide useful targets for interventions to reduce depression risk.

Adopting a biopsychosocial approach through incorporating a variety of mechanistic factors can capture the multidimensionality of the sleep-depression relationship, providing insight into the interplay across several systems. Although the effect size of our significant mediator was small, which is common when examining complex psychological phenomena (Götz, Gosling, & Rentfrow, 2022), and particularly in large population-based studies, it supports depression risk as being shaped by the combined impact of multiple biopsychosocial pathways. Identifying specific mechanisms linking adolescent sleep behaviours with depression in early adulthood, such as irritability, is necessary to inform practice, public health efforts, policy, and further research to generate a holistic understanding and effective tailoring of preventative efforts.

### Strengths and Limitations

This study has several strengths. First, the large sample size which is population-based and the longitudinal design. Second, involving young people with lived experience of mental health challenges to inform the selection of potential biopsychosocial mechanisms for our analyses. Third, our depression outcome was assessed using validated interviews. Fourth, adopting a holistic biopsychosocial approach to capture the multidimensionality of how sleep influences depression.

Nevertheless, there are some limitations. First, sleep was self-reported and assessed at a single time-point, which may be biased and not representative of one’s typical sleep behaviour. Therefore, future research examining how perceived and physiologically assessed WCS, SJL and sleep debt over several time-points relates with depression is warranted. Second, depression at 16 years was included as a covariate to account for prior symptom levels; however, as this was measured marginally post-exposure, the model may have underestimated the overall influence of sleep on subsequent depression (i.e., overadjustment bias) (Schisterman, Cole, & Platt, 2009). Third, our depression outcome variable included all participants who met criteria for a suggested diagnosis of mild, moderate, or severe depression, which may miss critical differences in disorder severity. Given the ALSPAC is population-based whereby the sample prevalence of severe depression at 24 years is low, merging severity levels maximised statistical power and still captured clinically relevant levels of depression. Future studies using clinical samples may be better suited to determine how sleep links with greater depression severity. Fourth, despite testing several theory- and PPIE-informed risk-enhancing and risk-reducing biopsychosocial factors, only those exerting a positive association with depression risk were significant in our data-driven selection process. Consequently, our path analysis was restricted to modelling risk-enhancing pathways with risk-enhancing sleep predictors (i.e., sleep debt). Therefore, we were unable to address potential protective mechanisms through which sleep may reduce depression risk. Future work focusing specifically on protective mediators, such as coping mechanisms, self-esteem and emotional stability (Tietbohl-Santos et al., 2024), is warranted. Finally, the significant indirect effect of irritability was small, thus only accounting for a small fraction of the association between sleep debt with later depression. This constrains the strength of the mechanistic conclusions, yet these findings are consistent with biopsychosocial models of depression whereby risk increases through the combined influence of several pathways (Remes, Mendes, & Templeton, 2021).

## Conclusion

In this longitudinal study, we found that weekday-weekend sleep duration inconsistency, specifically greater WCS in adolescence, and lower perceived sleep debt in adolescence was associated with lower depression risk in early adulthood. Furthermore, irritability emerged as a relevant biopsychosocial mechanism at 17 years, partially mediating the relationship between sleep debt and subsequent depression. WCS and sleep debt are common adolescent sleep behaviours which influence prospective depression in young adulthood. Promoting healthy sleep practices through ensuring adolescents have sufficient opportunity to increase their hours of sleep, via intervention strategies, may help protect against the development of future poor mental health.

## Availability of Data and Materials

The informed consent obtained from ALSPAC participants does not allow for the data to be made freely available through any third party maintained public repository. However, data used for this submission can be made available on request to the ALSPAC Executive. The ALSPAC data management plan describes in detail the policy regarding data sharing, which is through a system of managed open access. Full instructions for applying for data access can be found here: http://www.bristol.ac.uk/alspac/researchers/access/. The ALSPAC study website contains details of all the data that are available (http://www.bristol.ac.uk/alspac/researchers/our-data/).

M.P. and I.M.M had full access to all the data in the study and take responsibility for the integrity of the data and the accuracy of the data analysis. The scripts employed in generating the sleep variables and the analysis of the primary model are publicly available (https://osf.io/rkftd/).

## Data Availability

The informed consent obtained from ALSPAC participants does not allow for the data to be made freely available through any third party maintained public repository. However, data used for this submission can be made available on request to the ALSPAC Executive. The ALSPAC data management plan describes in detail the policy regarding data sharing, which is through a system of managed open access. Full instructions for applying for data access can be found here: http://www.bristol.ac.uk/alspac/researchers/access/. The ALSPAC study website contains details of all the data that are available (http://www.bristol.ac.uk/alspac/researchers/our-data/).
M.P. and I.M.M had full access to all the data in the study and take responsibility for the integrity of the data and the accuracy of the data analysis. The scripts employed in generating the sleep variables and the analysis of the primary model are publicly available (https://osf.io/rkftd/).

## Acknowledgements

We are incredibly grateful to all families who took part in this study, the midwives for their assistance in recruiting them, and the entire ALSPAC team, which includes interviewers, computer and laboratory technicians, clerical workers, research scientists, volunteers, managers, nurses, and receptionists.

