## Supplementary Information for "Adolescent Weekend Catch-Up Sleep and Sleep Sufficiency: Protective Factors for Depression in Young Adulthood"

**Figure S1.** *Flow chart of participation in the Avon Longitudinal Study of Parents and Children (ALSPAC)*

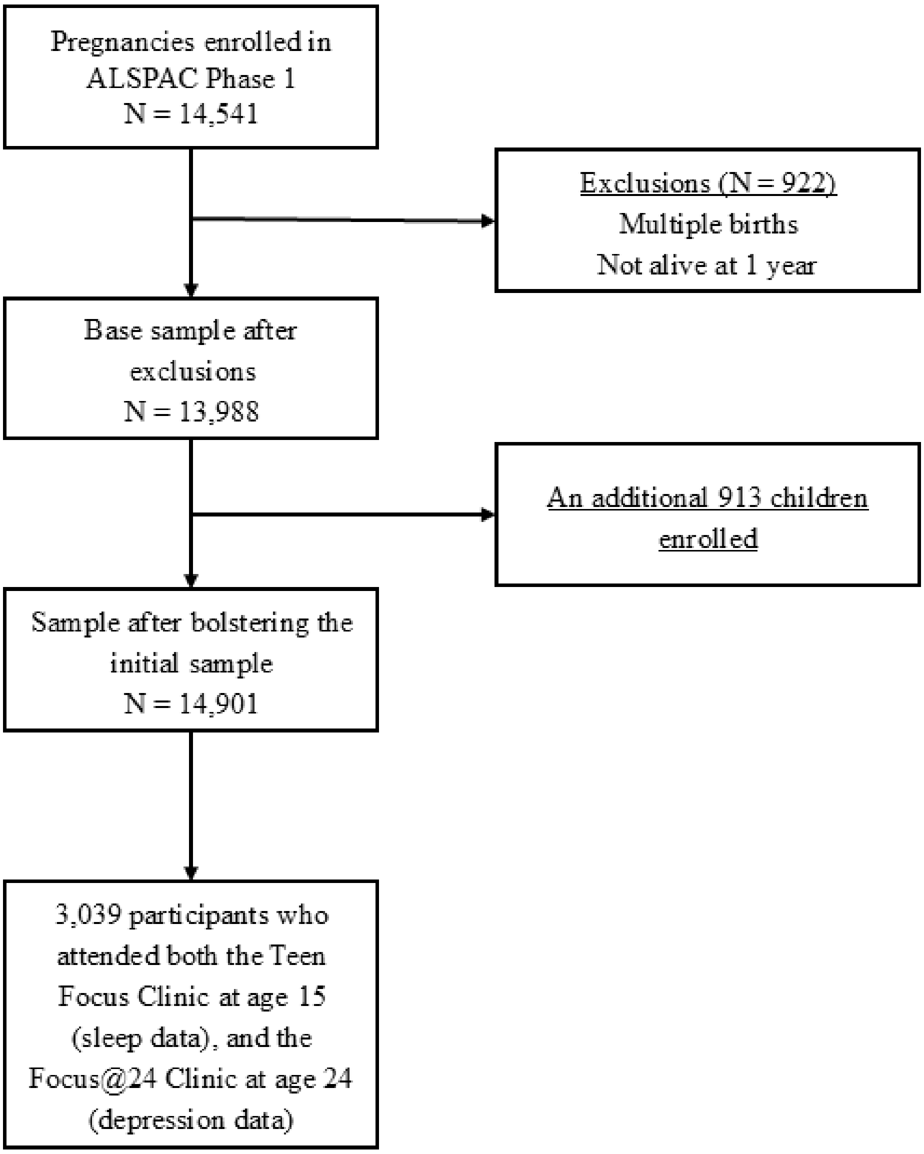


*Note.* ALSPAC: Avon Longitudinal Study of Parents and Children; N: Sample size.

**Appendix S1.** *Further information on the ALSPAC cohort*

The Avon Longitudinal Study of Parents and Children (ALSPAC), also known as the "Children of the 90s", is an ongoing population-based birth cohort established in the early 1990s in and around Bristol, United Kingdom to investigate how genetic, environmental, and social factors influence health and development across the life course (Boyd et al., 2013; Fraser et al., 2013). Pregnant women who resided in the former county of Avon with expected delivery dates between 1st April 1991 and 31st December 1992 were invited to participate, and 14,541 pregnancies were initially enrolled, defined as those with at least one returned questionnaire or attendance at a "Children in Focus" clinic by July 1999. These pregnancies resulted in 14,676 foetuses, 14,062 live births, and 13,988 children alive at one year of age. When the oldest cohort members were approximately seven years old, additional recruitment phases were employed to enrol eligible families who had not joined originally, increasing the baseline sample for analyses using data collected from age seven to age 24 to 15,454 pregnancies. The phases of enrolment are described in greater detail in the cohort profile papers and its update (Boyd et al., 2013; Fraser et al., 2013; Northstone et al., 2019). The total sample for analyses using any data collected after the age of seven is 15,447 pregnancies (15,658 foetuses). Amongst these, 14,901 children were alive at one year of age.

Study data were collected and managed using REDCap electronic data capture tools hosted at the University of Bristol. REDCap (Research Electronic Data Capture) is a secure, web-based software platform designed to support data capture for research studies (Harris et al., 2009).

Ethical approval for the study was obtained from the ALSPAC Ethics and Law Committee and the Local Research Ethics Committees. Informed consent for the use of all data collected was obtained from participants following the recommendations of the ALSPAC Ethics and Law Committee at the time. Participants can contact the study team at any time to retrospectively withdraw consent for their data to be used. Study participation is voluntary and during all data collection sweeps, information was provided on the intended use of data.

**Appendix S2.** *Further information on the selection of mechanistic factors*

Given the lack of studies examining the mechanistic role of biopsychosocial variables between our specific sleep variables and subsequent depression, we consulted the Youth Advisory Group (YAG) from the University of Birmingham's Institute for Mental Health. YAG members helped define which factors may be most relevant, which was then further clarified within the literature.

Prior to the focus group, all potential mediating factors were organised into overarching thematic categories identified by the researchers, including psychosis-related symptoms, common mental health symptoms, biological, alcohol/substance misuse, and school/work factors. Following focus group discussions, YAG members nominated three themes as the most relevant: common mental health symptoms, biological, and school/work factors. Next, the individual factors corresponding to these themes were presented for further review and consultation within the YAG group. This enabled identifying two specific factors from each theme that were considered most meaningful and appropriate to include in the analysis. This iterative collaboration with the YAG group aligns with ethical and methodological recommendations for conducting youth mental health research (Sales et al., 2021).

**Table S1.** *Overarching themes of factors*

| **Themes** | **Example subfactors** |
|---|---|
| Biological factors | Haematological markets, fatty acids, amino acids, etc. |
| Alcohol/substance misuse | Cannabis, cocaine, hallucinogens, etc. |
| School/work factors | In education/employment, connectedness to school/peers/colleagues, etc. |
| Mental health symptoms 1 (common mental health conditions) | Concentration, fatigue, irritability, etc. |
| Mental health symptoms 2 (psychosis-related symptoms) | Hallucinations, delusions, impact on daily life, etc. |

*Note.* Mental health symptoms 1 includes the symptom groups and their corresponding individual variables from the Clinical Interview Schedule-Revised (CIS-R). Mental health symptoms 2 includes items from the Psychosis-Like Symptoms Questionnaire (PLIKS) which is used to assess positive psychotic experiences in adolescents and young adults.

**Table S2.** *Selected factors and the measurements used*

| Factors | Measurement |
|---|---|
| Glucose | Calculated using the Homeostasis Model Assessment for Insulin Resistance ($HOMA_2$) computer-based formula which takes into account fasting plasma glucose and fasting insulin levels. These biochemical measurements were taken following an overnight fast or at least 6-8 hours prior to the clinic visit. $HOMA_2$ provides a continuous measure for glucose metabolism and is widely used to characterise various physiological subtypes of metabolic disorders. Scores are expressed as a percentage of a normal, healthy young adult whereby 100% (1.0) is considered normal function or sensitivity and increased scores indicate insulin resistance. |
| Cholesterol | Blood samples were taken following standard procedures with participants in a fasting state (asked to fast overnight or at least 6-8 hours prior to the clinic visit). The collected samples were immediately centrifuged and frozen at -80˚C. Total cholesterol, along with several other plasma lipid measures, was analysed according to the standard Lipid Research Clinics Protocol using enzymatic reagents for lipid determination. This study focused specifically on total cholesterol. |
| School connectedness | Self-reported item: "How well do you feel that you fit in at your school or college?". Response options included: "1 = Fit in very well", "2 = Fit in quite well", "3 = Some difficulty fitting in", "4 = Don't fit in at all", and "9 = Don't know". Cohort members were categorised as feeling connected to school if they fit in very or quite well through |

| | |
|---|---|
| | selecting response options “1” or “2”, whilst those who selected “3” or “4” were classified as not feeling connected. |
| Perceived academic difficulties | Self-reported item: “Do you have difficulty keeping up with your coursework or studies?”, with four response options: “1 = Never”, “2 = Occasionally”, “3 = Some of the time”, and “4 = Most of the time”. We then dichotomised this variable to reflect perceived academic difficulties if cohort members reported experiencing difficulties most of the time (“4”) and all other response choices reflected no academic difficulties (“1”, “2”, and “3”). |
| Fatigue | The fatigue total score (ranging from zero to four) from the CIS-R reflects the summed severity of fatigue-related symptoms over the past week, derived from items assessing persistent tiredness and low energy. Higher scores indicate more frequent and impairing fatigue symptoms. |
| Irritability | The irritability total score (ranging from zero to four) from the CIS-R represents the summed severity of irritability symptoms over the past week, based on items capturing a lowered threshold for annoyance, feeling easily irritated, and associated interference. Higher scores indicate more frequent and distressing irritability, reflecting heightened proneness to anger or annoyance in everyday situations. |

*Note.* CIS-R: Clinical Interview Schedule – Revised.

**Appendix S3.** *Supporting citations for biopsychosocial factors selected based on associations with depression and youth sleep outcomes*

- Homeostatic Model Assessment ($HOMA_2$) for Insulin Resistance (Al-Hakeim, Al-Kufi, Al-Dujaili, & Maes, 2018; Wilson, Carpenter, Song, Ho, & Hickie, 2021)
- Total cholesterol (Berentzen et al., 2014; Olguín-Montiel et al., 2025)
- School connectedness (Raniti, Rakesh, Patton, & Sawyer, 2022; Saletin et al., 2024)
- Perceived academic difficulties (Matos, Gaspar, Tomé, & Paiva, 2016; Zhang et al., 2022)
- Fatigue (Baum et al., 2014; Sunwoo, Kim, Chu, Yun, & Yang, 2022)
- Irritability (Baum et al., 2014; Hawes et al., 2020)

**Appendix S4.** *Further information on confounder selection*

Child's gestational age (weeks) (Pyhälä et al., 2017), recorded sex at birth (male versus female) (Daly, 2022), ethnicity (white versus other ethnic minority) (Flores, Sharp, Carson, & Cook, 2023), early family adversity (Wadman, Hiller, & St Clair, 2020), prior depression at 16 years (Johnson, Dupuis, Piche, Clayborne, & Colman, 2018), current sleep problems at 24 years (yes versus no) (Chunnan, Shaomei, & Wannian, 2022), and maternal age at birth (years) (Tearne et al., 2016), were included as covariates based on their association with depression.

Early family adversity was assessed using the Family Adversity Index (FAI) (Steer, Wolke, & Team, 2004) during pregnancy, at two and four years. Multiple dimensions (e.g., housing and family conditions, maternal education, financial challenges, etc.) were summed at each time point to generate a total FAI score, with higher scores indicating greater early family adversity.

Prior depression at 16 years was measured using the 13-item self-reported Short Mood and Feelings Questionnaire (SMFQ) whereby higher total scores (ranging from zero to 26) represent greater symptom severity over the previous two weeks (Angold et al., 1995). This was included to account for pre-existing depression levels.

Current sleep problems at 24 years were recorded as "Yes" or "No" in response to "In the PAST MONTH, have you been having problems with trying to get to sleep or with getting back to sleep if you woke up or were woken up?", which was part of the CIS-R (Lewis, Pelosi, Araya, & Dunn, 1992).

**Table S3.** *Associations between various sociodemographic variables possibly related to attrition and participation at 24 years of age*

| | Participation at 24 years | | |
|---|---|---|---|
| | **OR** | **95% CI** | ***P* value** |
| Sex | 0.468 | 0.435, 0.505 | <.001* |
| Child ethnicity | 1.417 | 1.168, 1.720 | <.001* |
| Birthweight, grams | 1.000 | 1.000, 1.000 | .003* |
| Maternal age at birth | 1.083 | 1.075, 1.092 | <.001* |
| Gestation | 1.092 | 1.078, 1.108 | <.001* |
| Family adversity index | 1.000 | 1.000, 1.000 | .501 |

*Note.* Logistic regression results with participation as an outcome and sociodemographic variables that have been related to systemic attrition bias as predictors. All variables except Family adversity index were significantly associated with participation at 24 years of age. OR: Odds ratio; CI: Confidence interval. *p<.05.

**Table S4.** *Number of participants with missing data for variables of interest among participants with complete outcome data*

| | Missing data, N (%) |
|---|---|
| **Sleep predictor variables at 15 years** | |
| Weekend catch-up sleep | 1208 (30.94%) |
| Social jetlag | 1208 (30.94%) |
| Sleep debt | 1253 (32.10%) |
| **Covariates** | |
| Child's gestational age | 285 (7.30%) |
| Maternal age when born | 285 (7.30%) |
| Early family adversity (FAI) | 400 (10.25%) |
| Depression (SMFQ), 16 years | 1197 (30.67%) |
| Ethnicity (White/Non-white) | 439 (11.24%) |
| Sex (Male/Female) | 2 (0.01%) |
| Sleep problems, 24 years (Present/No sleep problems) | 40 (1.02%) |
| **Mediating factors at 17 years** | |
| Glucose-insulin homeostasis ($HOMA_2$, index) | 2074 (67.03%) |
| Lipid homeostasis (Total cholesterol, mmol/L) | 1903 (48.74%) |
| School connectedness (Yes/No) | 1652 (42.32%) |
| Perceived academic difficulties (Yes/No) | 1638 (41.96%) |
| Fatigue (CIS-R subscale) | 1133 (29.02%) |
| Irritability (CIS-R subscale) | 1133 (29.02%) |

*Note.* FAI: Family Adversity Index; SMFQ: Short Moods and Feelings Questionnaire; $HOMA_2$: Homeostasis Model Assessment for Insulin Resistance; CIS-R: Clinical Interview Schedule – Revised.

**Table S5.** *Weighted adjusted associations between sleep variables at 15 and depression at 24 with covariates*

| | Adjusted Model (N=1899) | | |
|---|---|---|---|
| | **OR** | **95% CI** | ***P* value** |
| Sleep duration consistency at 15 years | 0.899 | 0.835, 0.967 | .004* |
| Social jetlag at 15 years | 0.933 | 0.827, 1.053 | .263 |
| Sleep debt at 15 years | 1.104 | 1.030, 1.184 | .005* |
| Sex | 0.964 | 0.799, 1.162 | .697 |
| Ethnicity | 1.279 | 0.841, 1.945 | .249 |
| Early family adversity | 1.063 | 1.040, 1.087 | <.001* |
| Child's gestational age | 1.029 | 0.980, 1.080 | .250 |
| Maternal age at birth | 0.976 | 0.956, 0.997 | .023* |
| Depression at 16 years | 1.103 | 1.087, 1.119 | <.001* |
| Sleep problems at 24 | 3.304 | 2.749, 3.969 | <.001* |

*Note.* OR: Odds ratio; CI: Confidence interval. *p<.05.

**Table S6.** *Weighted adjusted associations between biopsychosocial factors at 17 years and depression at 24 years*

| | Adjusted Model (N=988) | | |
|---|---|---|---|
| | **OR** | **95% CI** | ***P* value** |
| School connectedness | 0.853 | 0.561, 1.296 | .456 |
| Perceived academic difficulties | 1.742 | 1.181, 2.568 | .005* |
| Fatigue | 1.055 | 0.958, 1.161 | .275 |
| Irritability | 1.317 | 1.166, 1.489 | <.001* |
| Cholesterol | 1.512 | 1.231, 1.857 | <.001* |
| Glucose | 0.944 | 0.706, 1.261 | .695 |
| Sex | 0.790 | 0.582, 1.071 | .129 |
| Ethnicity | 2.447 | 1.157, 5.175 | .019* |
| Early family adversity | 1.098 | 1.058, 1.140 | <.001* |
| Child's gestational age | 1.020 | 0.943, 1.103 | .621 |
| Maternal age at birth | 0.986 | 0.955, 1.018 | .400 |
| Depression at 16 years | 1.070 | 1.045, 1.096 | <.001* |
| Sleep problems at 24 | 4.232 | 3.153, 5.679 | <.001* |

*Note.* OR: Odds ratio; CI: Confidence interval. *p<.05.

**Appendix S5.** *Path analysis model fit*

After inspecting global model fit, areas of local misfit which were reflected in the modification indices and standardised residuals covariances identified potential paths that aligned with both the theoretical framework as well as previous empirical studies.

Six further direct paths were sequentially added between covariates and the biopsychosocial mediators. Following each additional path, the model was re-estimated, and the global fit was re-checked. The table below summarises the sequence of modifications and associated fit indices for the overall model following each additional direct path.

**Table S7.** *Changes in overall model fit for path analysis model examining the mediating role of biopsychosocial factors between sleep debt and depression*

| **Added path** | $\chi^2$ | **RMSEA** | **CFI** |
|---|---|---|---|
| Depression → Irritability | 138.244; $p$<.001 | 0.032 | 0.979 |
| Depression → Perceived academic difficulties | 44.146; $p$<.001 | 0.019 | 0.994 |
| Depression → Cholesterol | 25.883; $p$<.001 | 0.015 | 0.997 |
| Early family adversity → Perceived academic difficulties | 16.355; $p$=.006 | 0.012 | 0.998 |
| Early family adversity → Cholesterol | 11.957; $p$=.018 | 0.011 | 0.999 |
| Maternal age at birth → Irritability | 6.258; $p$=.100 | 0.008 | 0.999 |

*Note.* $\chi^2$: Chi-square statistic; RMSEA: Root mean square error of approximation; CFI: Comparative fit index.
